# Comprehensive Thyroid Assessment in a Prospective Japanese Cohort: Epidemiology, Determinants, and Ultrasonographic Reference Ranges

**DOI:** 10.1101/2025.09.04.25335004

**Authors:** Satoko Shimazu-Kuwahara, Ichiro Yamuchi, Sachiko Kawashima, Makiko Tatsumi, Takuro Hakata, Yoriko Sakane, Masahiro Yakami, Kosuke Inoue, Daisuke Yabe, Mayumi Inoue

## Abstract

**Context:** A comprehensive evaluation of thyroid disease and health through multimodal assessment is warranted.

**Objective:** To clarify the epidemiology and clinical significance of abnormal findings on thyroid examinations in a medical checkup setting.

**Design:** Prospective cohort study conducted between April 1, 2016, and December 31, 2021.

**Setting:** Japanese adults undergoing self-paid medical checkups at the Preemptive Medicine and Lifestyle Disease Research Center, Kyoto University Hospital

**Main Outcome Measures:** All subjects underwent thyroid function tests, ultrasonography, and ^18^F-fluorodeoxyglucose-positron emission tomography (FDG-PET); anti-thyroperoxidase antibody (TPOAb) titers were measured in a subset of subjects.

**Results:** In the original cohort of 4,407 subjects (2,643 males and 1,764 females), the prevalence of thyroid dysfunction, increased blood flow on ultrasonography, diffuse thyroid FDG uptake, and thyroid nodules was 5.81%, 2.45%, 3.43%, and 39.71%, respectively; all were more frequent in females. Among 2,420 subjects with TPOAb measurements, TPOAb positivity was 7.19% and was significantly associated with thyroid dysfunction only at titers ≥ 128 IU/mL. Multivariate analyses identified age, sex, and thyroid volume as major determinants of thyroid function. Using data from 1,840 subjects without any thyroid abnormalities, we established sex-specific reference ranges for thyroid dimensions and found their correlations with age and body size.

**Conclusions:** This cohort provides epidemiological and physiological insights into thyroid health by integrating findings from thyroid function tests, ultrasonography, FDG-PET, and TPOAb measurements. Furthermore, the present study highlights the associations across abnormal findings, relationships within thyroid physiology, and the clinical relevance of high-titer TPOAb.

## Introduction

Thyroid diseases are common in the general population. Thyroid dysfunction is clinically important because, if left untreated, it can lead to multiple comorbidities. In the United Kingdom, the incidence of hypothyroidism is 0.41% per year in females and 0.06% per year in males, whereas that of hyperthyroidism is 0.08% per year in females and negligible in males (1). In the United States, the prevalence of overt and subclinical hypothyroidism is 0.3% and 4.3%, respectively, whereas that of overt and subclinical hyperthyroidism is 0.5% and 0.7% (2). When thyroid dysfunction is detected, its cause should be investigated using additional tests. Hashimoto’s disease, the most frequent cause of hypothyroidism, is diagnosed by detecting thyroid autoantibodies, such as anti-thyroperoxidase (TPOAb) and anti-thyroglobulin antibodies (TgAb). In the United States, TPOAb and TgAb positivity has been reported in 13.0% and 11.5% of adults, respectively (2).

Thyroid ultrasonography is a key imaging modality for the diagnosis of thyroid diseases. Diffuse hypervascularity on color Doppler, known as thyroid inferno, is a hallmark of Graves’ disease, although increased blood flow can also be observed in some cases of Hashimoto’s thyroiditis (3). In addition, ultrasonography is essential for the detailed evaluation of thyroid nodules. The prevalence of thyroid nodules is high and increases with age, reaching nearly 50% when assessed using ultrasonography (4). ^18^F-fluorodeoxyglucose (FDG) positron emission tomography (PET) is widely used in oncological evaluations, and the thyroid gland occasionally exhibits FDG uptake. Diffuse thyroid FDG uptake suggests a high likelihood of Hashimoto’s disease (5,6). Although the prevalence of diffuse thyroid FDG uptake has been reported (5,7), its clinical significance remains unclear.

Although thyroid diseases can be evaluated using multiple modalities, data on the relationships among these examinations are limited, particularly regarding ultrasonography and FDG-PET. The Fukushima Health Management Survey reported estimated thyroid volume by ultrasonography from infancy through adolescence (8), whereas fewer data are available for adults (9,10). Moreover, in adults, the high prevalence of Hashimoto’s thyroiditis and thyroid nodules complicates the identification of truly disease-free reference populations. Therefore, a cohort restricted to individuals with healthy thyroid glands is warranted to investigate thyroid physiology.

In 2016, we launched a prospective cohort study of Japanese adults in a medical checkup setting. This cohort featured comprehensive phenotyping. For thyroid assessment, all subjects underwent thyroid function tests, thyroid ultrasonography, and FDG-PET; measurement of TPOAb was incorporated midway through the study. We aimed to clarify the epidemiology and clinical significance of abnormal findings detected in these thyroid examinations, focusing on the baseline cross-sectional dataset.

## Methods

### Subjects

A flow diagram of the enrollment process is shown in Figure 1. From April 1, 2016, to December 31, 2021, we enrolled 4,651 adults who underwent their initial comprehensive self-paid medical checkups at the Preemptive Medicine and Lifestyle Disease Research Center, Kyoto University Hospital. We excluded 5 individuals without blood tests and 111 who did not provide informed consent. In addition, 128 individuals were excluded because of current or past thyroid diseases, including Graves’ disease (n = 37), levothyroxine replacement for hypothyroidism (n = 37), thyroid cancer (n = 36), surgery for thyroid nodules (n = 13), and a history of subacute thyroiditis (n = 5). A total of 4,407 subjects met the inclusion criteria for this study, and we defined this group as the Original cohort (Figure 1). The study was approved by the Ethics Committee of Kyoto University Hospital (approval numbers R0619 and R3604), and written informed consent was obtained from all subjects.

**Figure 1.**
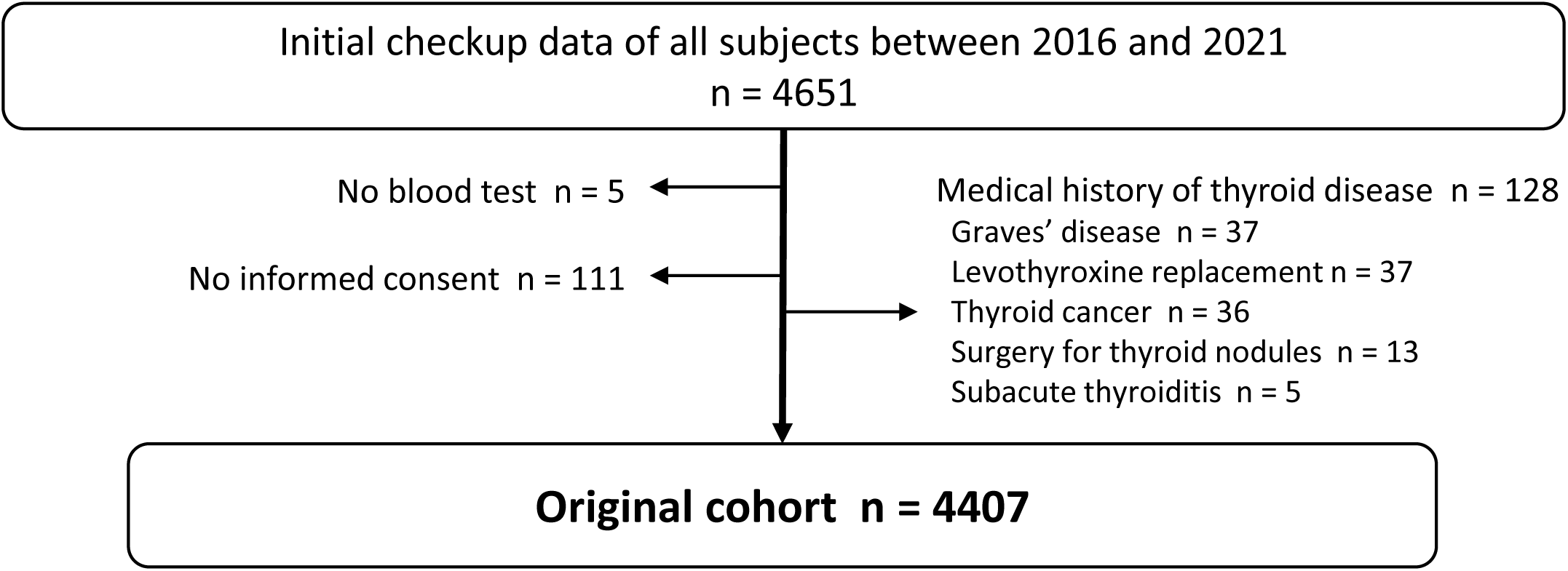
Flowchart illustrating the establishment of the Original cohort in this prospective study. n, number of subjects.

### Evaluation

Serum levels of TSH, free T4 (fT4), and free T3 (fT3) were measured using electrochemiluminescence immunoassays (Elecsys; Roche Diagnostics, Mannheim, Germany). Reference ranges were as follows: TSH, 0.500–5.000 μIU/mL; fT4, 0.880–1.620 ng/dL; and fT3, 2.33–4.00 pg/mL. TPOAb measurements were initiated on January 1, 2018, and were performed using an Elecsys assay (reference: < 16 IU/L). Thyroid ultrasonography was performed using an Aplio 500 system (Canon Medical System Corporation, Otawara, Japan) equipped with a linear transducer (PLT-1005BT; 6-cm footprint). We measured the isthmus thickness and, for each lobe, the transverse, anteroposterior, and longitudinal diameters. Lobe volume was calculated as the transverse diameter × anteroposterior diameter × longitudinal diameter × π/6, and total thyroid volume was obtained by summing volumes of both lobes. Blood flow was evaluated using color Doppler ultrasonography. The entire gland was systematically surveyed for nodules and cysts. Ultrasound cine loops were reviewed by a second sonographer for quality assurance. Increased blood flow was defined as positive only when both sonographers and a board-certified endocrinologist independently concurred. For FDG-PET, we focused on diffuse thyroid uptake and did not analyze focal uptake confined to nodules. FDG-PET images were independently reviewed by four board-certified radiologists, and uptake was considered positive only when multiple radiologists independently agreed on a distinct uptake.

### Statistical analyses

All statistical analyses were performed using JMP Pro version 18.0 (SAS Institute Inc., NC, USA). Unless otherwise specified, values are presented as the mean ± SD. As TSH values followed a log-normal distribution, statistical analyses were performed using log-transformed values. Continuous variables were compared using Student’s *t*-test, and categorical variables using Fisher’s exact test. For trend analyses, we assigned the median value of each ordered category and modeled it as a continuous variable in regression analyses. Multivariable linear regression was used to assess independent associations. Pearson’s correlation coefficients were calculated to evaluate correlations between variables. Significant differences were considered as *p* < 0.05.

## Results

### Prevalence of abnormal findings on thyroid examinations

The characteristics of the Original cohort are summarized in Table 1. The mean age was 52.9 ± 11.3 years; 2,643 (59.97%) were males and 1,764 (40.03%) were females. The serum levels of TSH, fT4, and fT3 were 1.952 ± 2.245 μIU/mL, 1.288 ± 0.200 ng/dL, and 3.08 ± 0.58 pg/mL, respectively. Significant sex-related differences were observed (Table 1). Males were slightly older and had larger anthropometric measurements than females. TSH levels were slightly lower in males than in females (1.859 vs. 2.091 μIU/mL, *p* = 0.0001), whereas fT4 and fT3 levels were higher in males (fT4, 1.313 vs. 1.250 ng/dL, *p* < 0.0001; fT3, 3.19 vs. 2.93 pg/mL, *p* < 0.0001).

**Table 1.** Characteristics and prevalence of abnormal findings in the Original cohort.

|  | All (n = 4407) | Male (n = 2643) | Female (n = 1764) | *p* |
| --- | --- | --- | --- | --- |
| Age (year) | 52.9 (11.3) | 53.2 (11.0) | 52.3 (11.9) | 0.0114 |
| Height (cm) | 166.2 (8.7) | 171.5 (6.1) | 158.3 (5.5) | < 0.0001 |
| Body weight (kg) | 66.0 (14.1) | 73.5 (11.5) | 54.7 (9.3) | < 0.0001 |
| BMI (kg/m<sup>2</sup>) | 23.7 (3.8) | 25.0 (3.5) | 21.8 (3.6) | < 0.0001 |
| Thyroid function |  |  |  |  |
| TSH (μIU/mL) | 1.952 (2.245) | 1.859 (1.972) | 2.091 (2.596) | 0.0001 |
| ft4 (ng/dL) | 1.288 (0.200) | 1.313 (0.180) | 1.250 (0.221) | < 0.0001 |
| ft3 (pg/mL) | 3.08 (0.58) | 3.19 (0.39) | 2.93 (0.76) | < 0.0001 |
| Thyroid dysfunction |  |  |  |  |
| No n (%) | 4151 (94.19%) | 2505 (94.78%) | 1646 (93.31%) | 0.0485 |
| Yes n (%) | 256 (5.81%) | 138 (5.22%) | 118 (6.69%) |  |
| Overt thyrotoxicosis | 27 (0.61%) | 15 (0.57%) | 12 (0.68%) |  |
| Subclinical thyrotoxicosis | 104 (2.36%) | 69 (2.61%) | 35 (1.98%) |  |
| Subclinical hypothyroidism | 118 (2.68%) | 52 (1.97%) | 66 (3.74%) |  |
| Overt hypothyroidism | 7 (0.16%) | 2 (0.08%) | 5 (0.28%) |  |
| Increased blood flow |  |  |  |  |
| No n (%) | 4299 (97.55%) | 2608 (98.68%) | 1691 (95.86%) | < 0.0001 |
| Yes n (%) | 108 (2.45%) | 35 (1.32%) | 73 (4.14%) |  |
| Diffuse FDG uptake |  |  |  |  |
| No n (%) | 4256 (96.57%) | 2604 (98.52%) | 1652 (93.65%) | < 0.0001 |
| Yes n (%) | 151 (3.43%) | 39 (1.48%) | 112 (6.35%) |  |
| Thyroid nodule |  |  |  |  |
| No n (%) | 2657 (60.29%) | 1763 (66.71%) | 894 (50.68%) | < 0.0001 |
| Yes n (%) | 1750 (39.71%) | 880 (33.29%) | 870 (49.32%) |  |
| < 5.0 mm | 667 (15.14%) | 393 (14.87%) | 274 (15.53%) |  |
| 5.0–9.9 mm | 705 (16.00%) | 329 (12.45%) | 376 (21.32%) |  |
| 10.0–19.9 mm | 315 (7.15%) | 133 (5.03%) | 182 (10.32%) |  |
| ≥ 20.0 mm | 63 (1.43%) | 25 (0.95%) | 38 (2.15%) |  |

BMI, body mass index; fT4, free T4; fT3, free T3; FDG, <sup>18</sup>F-fluorodeoxyglucose. Continuous variables are expressed as mean (SD). Comparisons between males and females were performed using the Student's *t*-test or Fisher's exact test, as appropriate. TSH values were log-transformed for statistical analyses due to their log-normal distribution.

Thyroid dysfunction was identified in 256 subjects (5.81%) (Figure 2A, Table 1). The most common pattern was subclinical hypothyroidism (n = 118; 2.68%), followed by subclinical thyrotoxicosis (n = 104; 2.36%), overt thyrotoxicosis (n = 27; 0.61%), and overt hypothyroidism (n = 7; 0.16%) (Figure 2B, Table 1). The characteristics of each group, categorized by thyroid dysfunction pattern, are shown in Table 2. Overt thyrotoxicosis was more common in younger subjects, whereas subclinical hypothyroidism predominated among older subjects and was more frequent in females.

**Figure 2.**
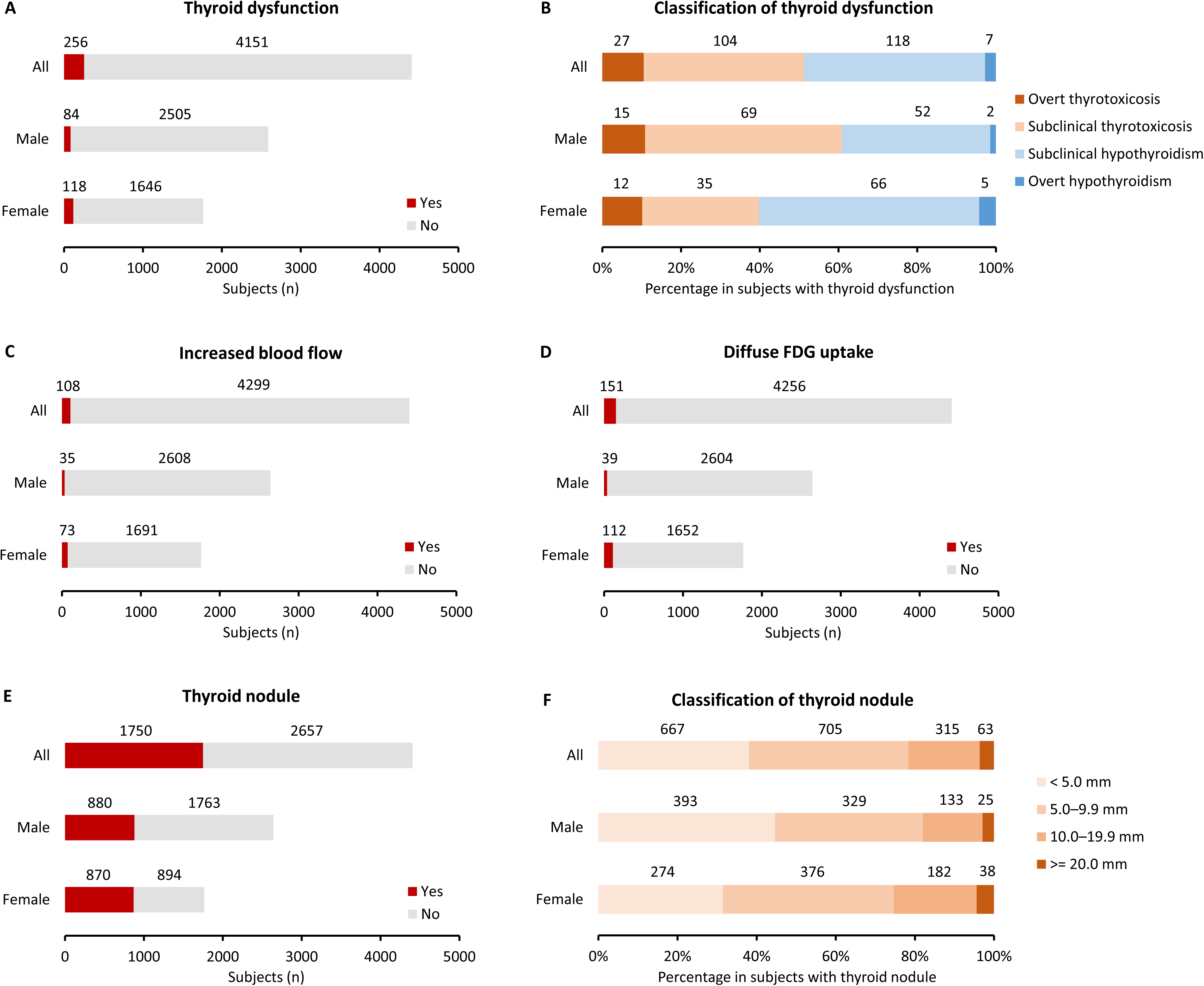
Prevalence of abnormal findings on thyroid examinations in the Original cohort. The actual number of subjects is shown in panels A, C, D, and E. The percentage of subjects with abnormal findings is shown in panels B and F. Thyroid nodules were classified by maximum diameter. FDG: ^18^F-fluorodeoxyglucose.

**Table 2.**
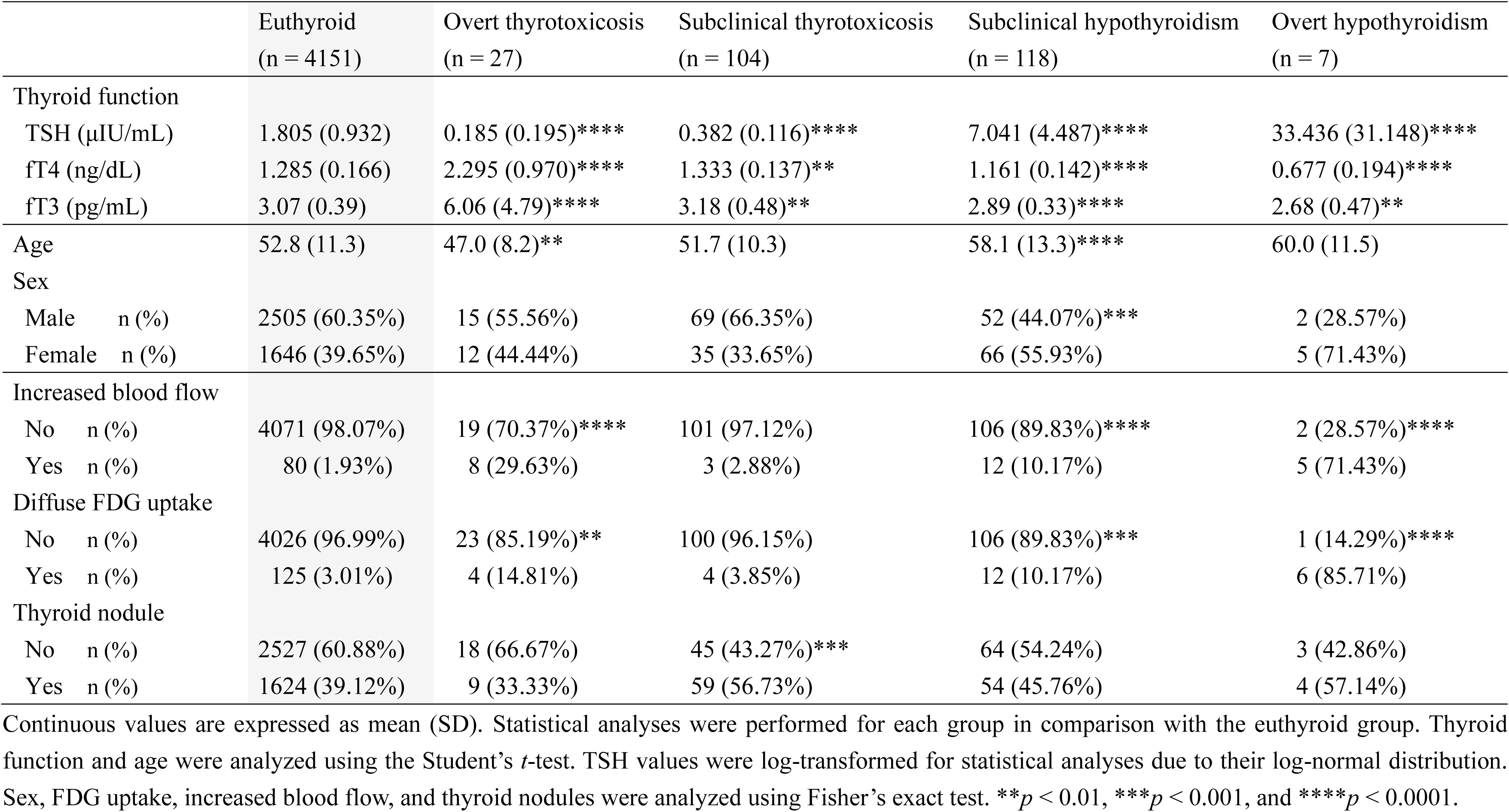
Characteristics and prevalence of abnormal findings in the Original cohort, classified by thyroid dysfunction.

Increased blood flow on thyroid ultrasonography was detected in 108 subjects (2.45%) and was significantly more common in females (4.14%) than in males (1.32%) (Figure 2C, Table 1). Diffuse thyroid FDG uptake was observed in 151 subjects (3.43%) and was likewise more common in females (6.35%) than in males (1.48%) (Figure 2D, Table 1). Both increased blood flow and diffuse FDG uptake were often present in the subjects with overt thyroid dysfunction or subclinical hypothyroidism, especially those with overt hypothyroidism (Table 2). Thyroid nodules were found in 1,750 subjects (39.71%) (Figure 2E, Table 1). Based on the maximum diameter, 667 subjects (15.14%) had nodules < 5.0 mm, 705 (16.00%) had nodules 5.0–9.9 mm, 315 (7.15%) had nodules 10.0–19.9 mm, and 63 (1.43%) had nodules ≥ 20.0 mm (Figure 2F, Table 1). Females had a higher prevalence of nodules ≥ 5.0 mm than males (33.79% vs. 18.43%) (Table 1). The prevalence of nodules increased with age, whereas the prevalence of thyroid dysfunction, increased blood flow, and diffuse FDG uptake did not show significant age-related variations (Supplementary Table 1).

### Association of clinical characteristics and abnormal findings with thyroid function

We subsequently assessed the relationship of clinical characteristics and abnormal findings with thyroid function. Although fT4 levels showed a significant correlation with log-transformed TSH levels (r = −0.3829, *p* < 0.0001) (Figure 3A), we excluded subjects whose thyroid function values were outside the main distribution for subsequent analyses (Figure 3B): those with TSH < 0.1 μIU/mL (n = 18), TSH > 10 μIU/mL (n = 13), and one subject with syndrome of inappropriate secretion of TSH (SITSH, n = 1). In this dataset restricted to subnormal thyroid function, older age was associated with higher TSH levels in both sexes (Figure 3C, Supplementary Table 2). Sex differences were observed in fT4 and fT3 trends: in males, both hormones decreased with age, whereas in females, only a slight age-related decline in fT3 was observed (Figure 3D and 3E, Supplementary Table 2). Height showed only a minor association with thyroid function, whereas body weight correlated positively with fT3 levels (Supplementary Figure 1, Supplementary Table 3 and 4). Body mass index (BMI) analysis showed that lean subjects had lower fT3 levels and obese subjects had higher fT3 levels in both sexes; no BMI-related trends were observed for TSH or fT4 (Figure 3F–3H, Supplementary Table 5).

**Figure 3.**
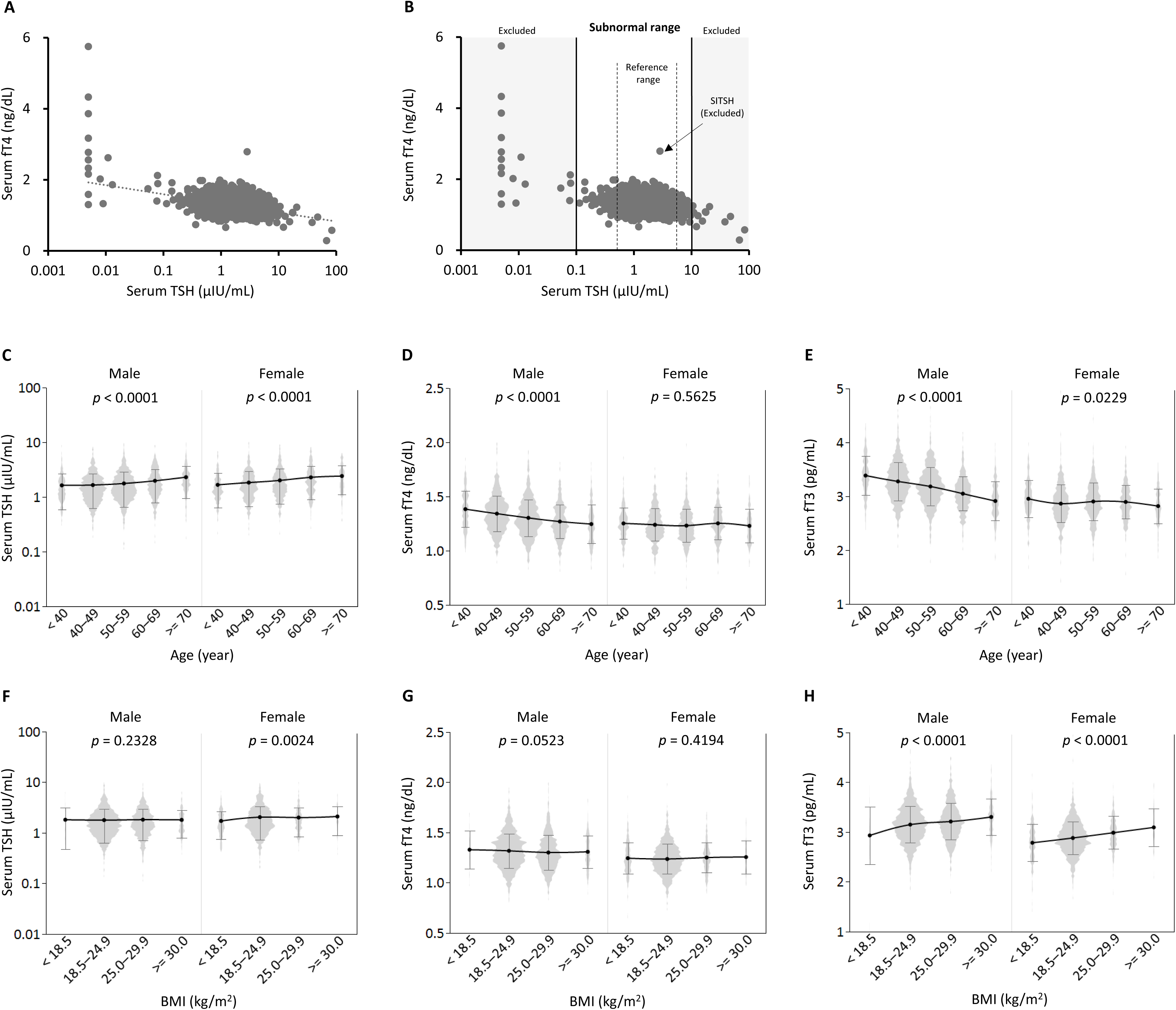
Associations between characteristics and thyroid function in the Original cohort. (A) Scatter plot of serum TSH and free T4 (fT4) levels. (B) Illustration of the exclusion criteria for the analyses presented in Figures 3 and 4, overlaid on a scatter plot. (C–E) Trends in thyroid function according to age (see Supplementary Table 2 for the actual values). (F–H) Trends in thyroid function according to body mass index (BMI) (see Supplementary Table 5 for actual values). SITSH, syndrome of inappropriate secretion of TSH. Gray violin plots show the distribution of the data, and black circles with connecting lines represent mean ± SD. Log-transformed TSH values were used for statistical analyses because of their log-normal distribution. Trend analyses were conducted by assigning the median value of each category and modeling it as a continuous variable in regression analyses.

We then examined the relationship between abnormal findings and thyroid function. Subjects with increased blood flow had higher TSH and lower fT4 levels, with no changes in fT3 levels (Figure 4A–4C, Supplementary Table 6). Diffuse FDG uptake was also associated with higher TSH and lower fT4 levels (Figure 4D–4F, Supplementary Table 6). Regarding thyroid nodules, only minor size-related trends were observed; nodules ≥ 20 mm were associated with lower TSH levels in females (Figure 4G–4I, Supplementary Table 7). Moreover, the estimated thyroid volume was analyzed using ultrasonography. Due to technical limitations, mainly the difficulty in measuring the longitudinal diameter, data were unavailable for 183 subjects, leaving 4,192 evaluable cases. Thyroid volume was negatively correlated with TSH and positively correlated with both fT4 and fT3 levels (Figure 4J–4L, Supplementary Table 8).

**Figure 4.**
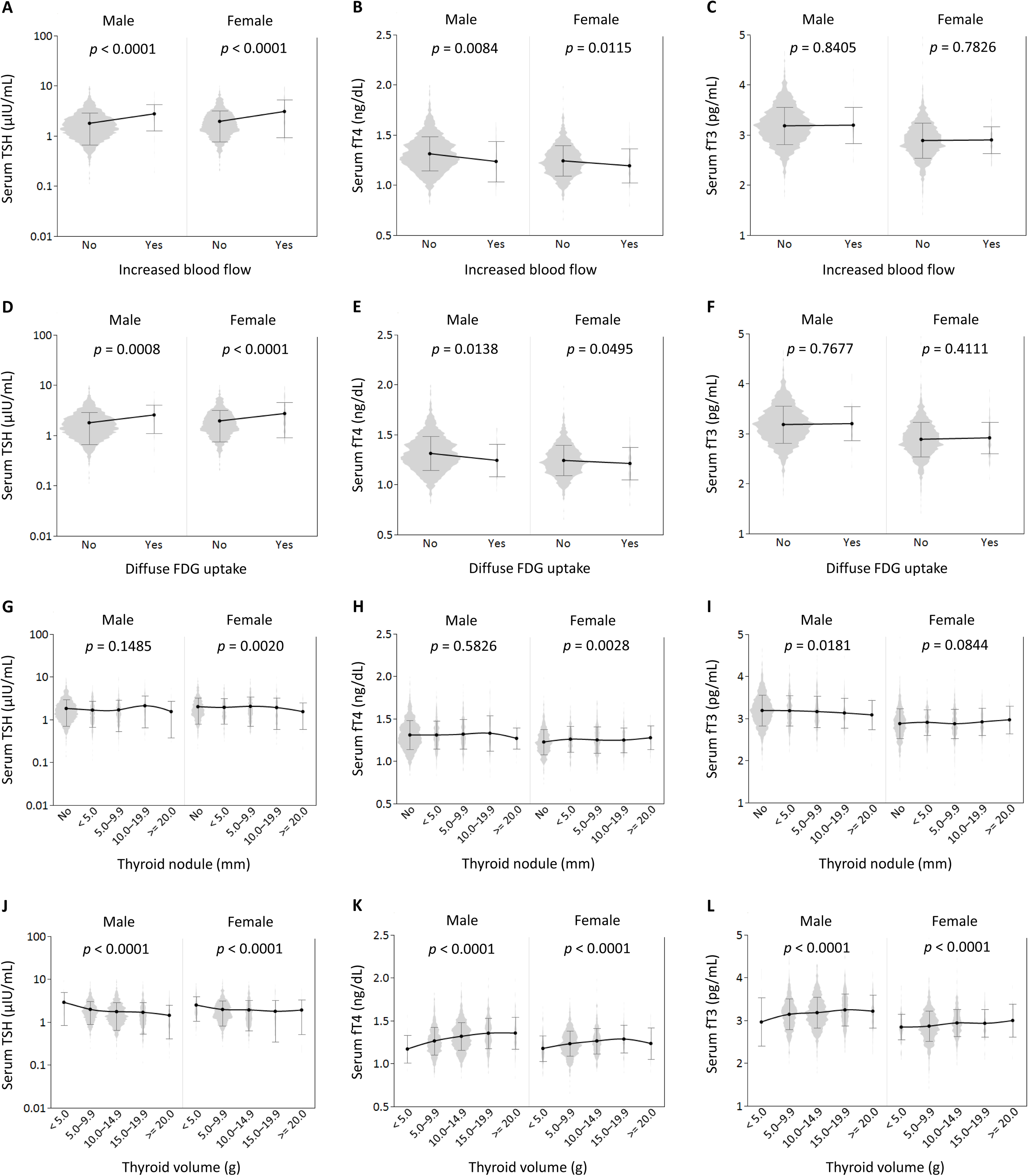
Associations between thyroid examination results and thyroid function in the Original cohort. (A–C) Differences in thyroid function according to the presence of increased blood flow on thyroid ultrasonography (see Supplementary Table 6 for actual values). (D–F) Differences in thyroid function according to the presence of diffuse FDG uptake (see Supplementary Table 6 for actual values). (G–I) Trends in thyroid function according to thyroid nodule size, classified by maximum diameter (see Supplementary Table 7 for the actual values). (J–L) Trends in thyroid function according to thyroid volume (see Supplementary Table 8 for actual values). Data from 183 subjects were excluded because of technical issues in volume measurement using ultrasonography. Gray violin plots show the distribution of the data, and black circles with connecting lines represent mean ± SD. Student’s *t* test was used for panels A–F. Trend analyses were performed for panels G–L.

### Association between TPOAb titers and thyroid function

As TPOAb measurements began partway through the study, we additionally analyzed a TPOAb cohort consisting of 2,420 subjects with available TPOAb results (Figure 5A). The characteristics of this cohort were similar to those of the Original cohort (Supplementary Table 9). TPOAb positivity was found in 174 subjects (7.19%), with a higher prevalence in females (96 of 938 subjects, 10.23%) than in males (78 of 1,482 subjects, 5.26%; p < 0.0001) (Figure 5B). Subjects with TPOAb positivity more frequently exhibited abnormal findings on thyroid examinations than those without TPOAb; thyroid dysfunction (13.22% vs. 4.94%, p < 0.0001), increased blood flow (28.16% vs. 1.11%, p < 0.0001), and diffuse FDG uptake (28.74% vs. 1.25%, p < 0.0001) (Figure 5C–5E, Supplementary Table 10). The prevalence of thyroid nodules did not differ between TPOAb-positive and TPOAb-negative subjects (40.80% vs. 39.45%, p = 0.7478) (Figure 5F, Supplementary Table 10).

**Figure 5.**
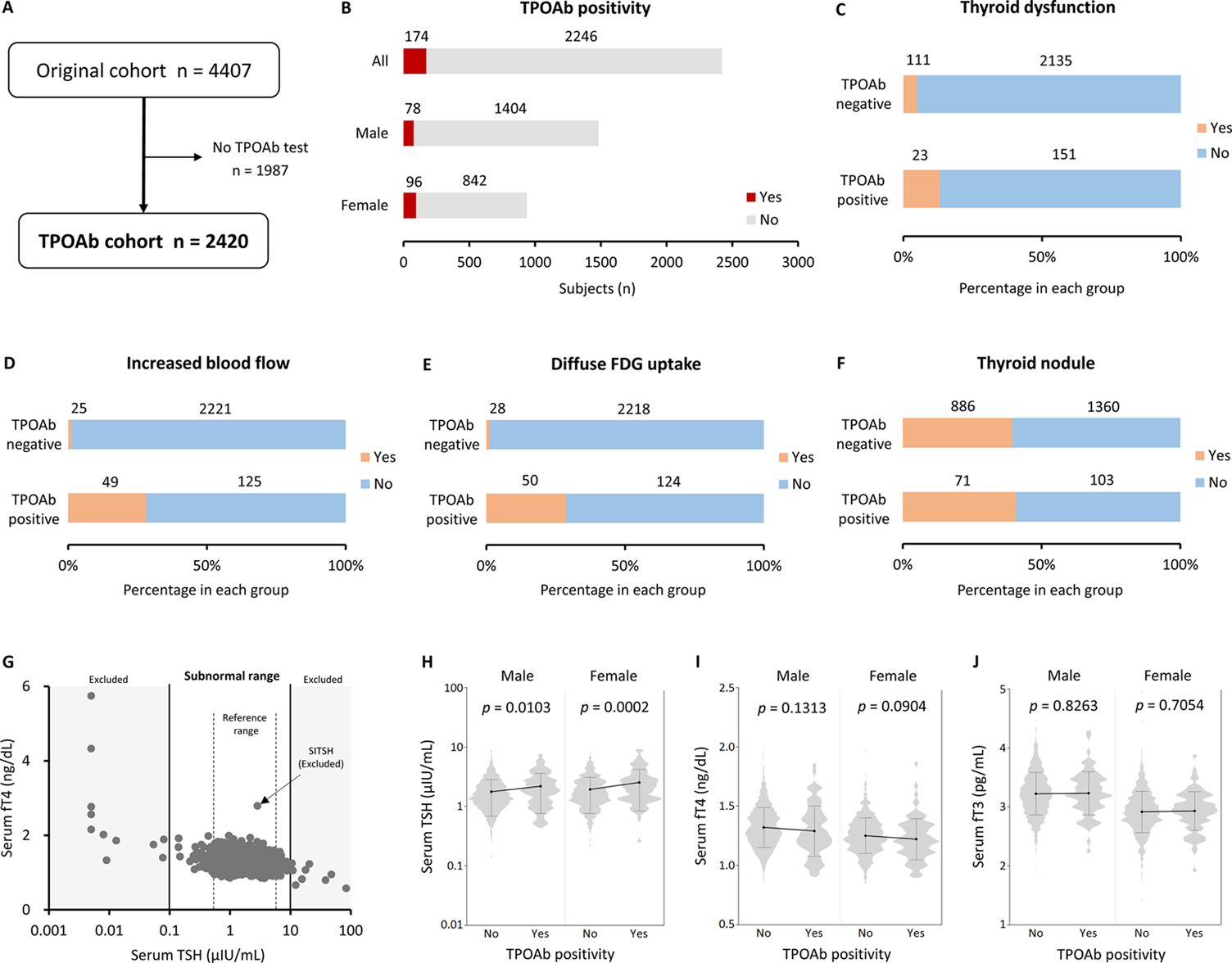
Associations between TPOAb titers and thyroid examination results in the TPOAb cohort. (A) Flowchart illustrating the selection of the TPOAb cohort. (B) Number of TPOAb-positive subjects. (C–F) Prevalence of abnormal findings in TPOAb-positive subjects; any thyroid dysfunction (C), increased blood flow (D), diffuse FDG uptake (E), and any thyroid nodules (F). (G) Exclusion criteria for analyses in panels H–J overlaid on the scatter plot. (H–J) Differences in thyroid function according to TPOAb positivity (see Supplementary Table 11 for the actual values). TPOAb, anti-thyroperoxidase antibody. Gray violin plots show the distribution of the data, and black circles with connecting lines represent mean ± SD. Student’s *t* test was used.

We subsequently examined whether the TPOAb titer influenced these associations. Thyroid dysfunction, particularly hypothyroidism, was more frequent among subjects with TPOAb titers ≥ 128 IU/mL (Table 3). The prevalence of increased blood flow and diffuse FDG uptake also increased in a titer-dependent manner (Table 3). To further clarify the relationship between TPOAb positivity and thyroid function, we excluded subjects whose thyroid function values were outside the main distribution (Figure 5G), as in Figure 3B: TSH < 0.1 μIU/mL (n = 11), TSH > 10 μIU/mL (n = 10), or SITSH (n = 1). In this dataset, TPOAb-positive subjects had higher TSH levels than TPOAb-negative subjects, whereas fT4 and fT3 levels remained unchanged (Figure 5H–5J, Supplementary Table 11). Analysis by titer showed that TSH elevation was observed only in subjects with TPOAb ≥ 128 IU/mL, with no corresponding changes in fT4 or fT3 levels (Supplementary Figure 2, Supplementary Table 11).

**Table 3.**
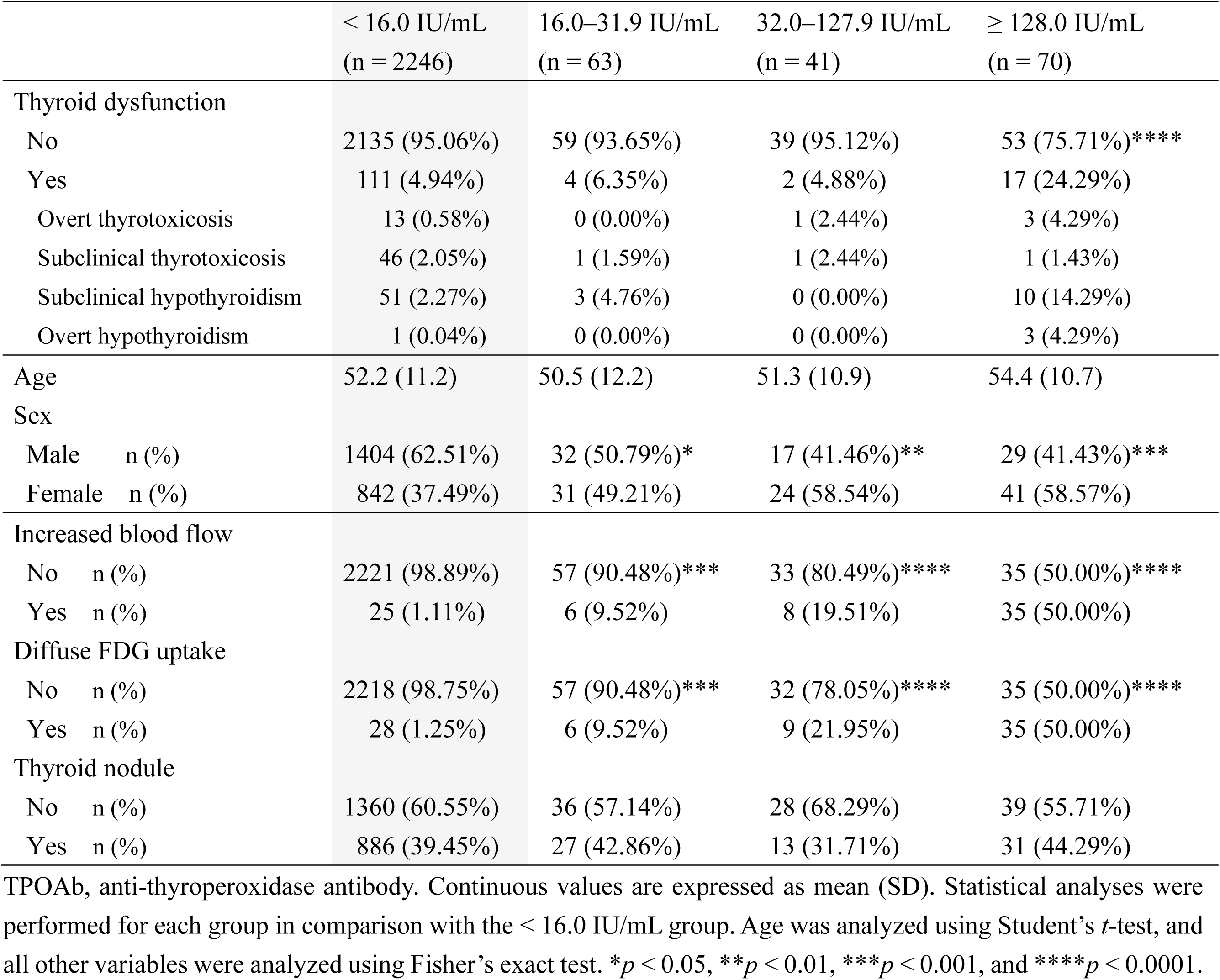
Characteristics and prevalence of abnormal findings in the TPOAb cohort, classified by TPOAb titers.

From these analyses, we identified age, sex, BMI, increased blood flow, diffuse FDG uptake, thyroid volume, and TPOAb titers ≥ 128 IU/mL as potential determinants of thyroid function. To assess their relative contributions, we conducted multivariable analyses within the TPOAb cohort after excluding subjects lacking thyroid volume data (n = 130), those with nodules ≥ 20 mm, and those with outlying thyroid function values (TSH < 0.1 μIU/mL, n = 6; TSH > 10 μIU/mL, n = 10; SITSH, n = 1). Among the 2,239 subjects analyzed, Model 1, which included only demographic and anthropometric variables, showed an age-related increase in TSH and decreases in fT4 and fT3 levels. Sex remained a significant determinant even after adjusting for age and BMI: males had lower TSH and higher fT4 and fT3 levels than females. BMI showed a positive correlation exclusively with fT3 levels (Table 4). Model 2, which incorporated all identified factors, reproduced the age-related changes, but the association between TSH levels and sex was no longer significant. BMI was weakly associated with TSH and fT3 levels in both sexes. Among the factors obtained from thyroid examinations, thyroid volume was negatively associated with TSH and positively associated with fT4 and fT3 levels; increased blood flow and TPOAb ≥ 128 IU/mL were linked to higher TSH levels; and increased blood flow and diffuse FDG uptake were associated with lower fT4 levels. In summary, age and sex remain the dominant determinants of thyroid function, while ultrasonographic findings have a significant influence.

**Table 4.** Multivariate regression analysis for determinants of thyroid function.

|  | Model 1 |  |  | Model 2 |  |  |
| --- | --- | --- | --- | --- | --- | --- |
|  | logTSH | fT4 | fT3 | logTSH | fT4 | fT3 |
| **R<sup>2</sup>** | 0.039 | 0.062 | 0.209 | 0.096 | 0.107 | 0.212 |
| **Intercept** | −0.023 | 1.362 | 3.036 | 0.199 | 1.257 | 3.016 |
| Age (year) | 0.004**** | −0.002**** | −0.007**** | 0.004**** | −0.002**** | −0.007**** |
| Sex (male to female) | −0.023*** | 0.034**** | 0.128**** | 0.002 | 0.020**** | 0.122**** |
| BMI (kg/m<sup>2</sup>) | 0.001 | 0.001 | 0.017**** | 0.005** | −0.001 | 0.016**** |
| Increased blood flow |  |  |  | 0.063** | −0.037** | −0.018 |
| Diffuse FDG uptake |  |  |  | 0.026 | −0.031* | 0.007 |
| Thyroid volume (g) |  |  |  | −0.014**** | 0.009**** | 0.005* |
| TPOAb ≥ 128.0 IU/mL |  |  |  | 0.062*** | 0.009 | 0.019 |

R<sup>2</sup>: coefficient of determination. Results are represented as estimates. "log" denotes the base-10 logarithm.
\* $p < 0.05$ , \*\* $p < 0.01$ , \*\*\* $p < 0.001$ , and \*\*\*\* $p < 0.0001$ .

### Reference ranges and factors associated with thyroid size

A major strength of this study is the strict definition of subjects without thyroid abnormalities based on multimodal thyroid assessment. Accordingly, we established a Thyroid-healthy cohort by excluding subjects with abnormal TSH levels, TPOAb positivity, increased blood flow, diffuse FDG uptake, or thyroid nodules (Figure 6A). The characteristics of the Thyroid-healthy cohort (n = 1,840) are presented in Table 5. Subjects with nodules < 10 mm were included, given their high prevalence (33.37%). All thyroid size parameters, including the isthmus thickness, the transverse, anteroposterior, and longitudinal diameters of each lobe, were larger in males than in females (Table 5). Several sex-independent correlations were observed: the longitudinal diameter was negatively correlated with age (Figure 6B–6E, Supplementary Table 12) and positively correlated with height (Figure 6F–6I, Supplementary Table 13). Body weight and BMI were positively correlated with the isthmus thickness and the transverse and anteroposterior diameters (Figure 6J–6M, Supplementary Figure 3, Supplementary Table 14 and 15). Consequently, thyroid volume was positively correlated with height, body weight, and BMI (Supplementary Figure 4, Supplementary Table 12–15).

**Figure 6.**
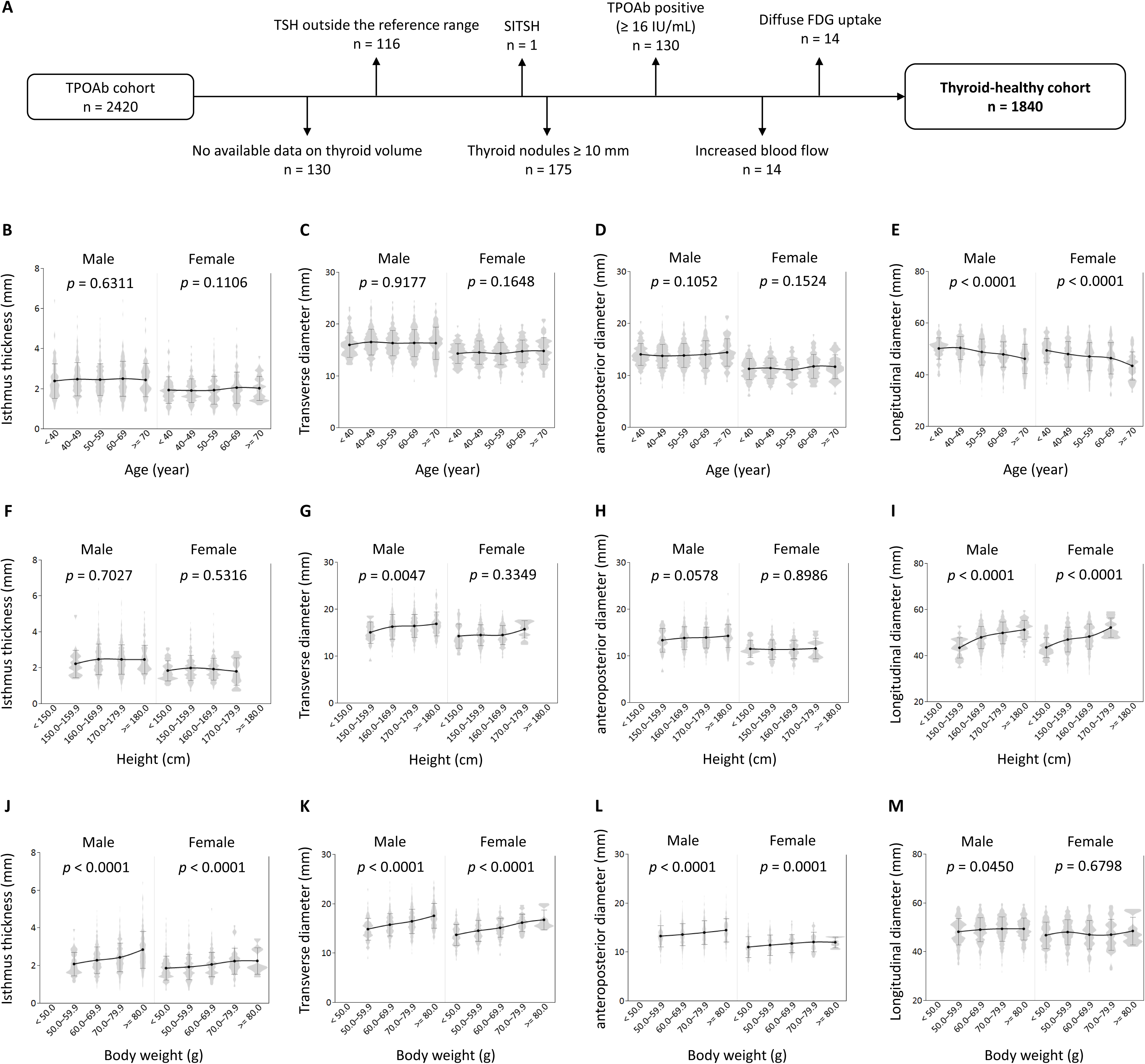
Associations between characteristics and thyroid size in the Thyroid-healthy cohort. (A) Flowchart illustrating the selection of the Thyroid-healthy cohort. (B–E) Trends in thyroid size according to age (see Supplementary Table 12 for the actual values). (F–I) Trends in thyroid size according to height (see Supplementary Table 13 for the actual values). (J–M) Trends in thyroid size according to body weight (see Supplementary Table 14 for actual values). Two male subjects weighing < 50.0 kg were excluded because of the small sample size. Gray violin plots show the distribution of the data, and black circles with connecting lines represent mean ± SD.

**Table 5.** Characteristics and thyroid size on ultrasonography in the Thyroid-healthy cohort.

|  | All (n = 1840) | Male (n = 1187) | Female (n = 653) | *p* |
| --- | --- | --- | --- | --- |
| Age | 51.7 (10.9) | 52.1 (10.6) | 51.1 (11.5) | 0.0780 |
| Height (cm) | 167.1 (8.5) | 171.8 (5.8) | 158.4 (5.5) | < 0.0001 |
| Weight (kg) | 67.3 (14.1) | 74.0 (11.6) | 55.0 (9.3) | < 0.0001 |
| BMI (kg/m<sup>2</sup>) | 23.9 (3.9) | 25.1 (3.5) | 21.9 (3.6) | < 0.0001 |
| Thyroid function |  |  |  |  |
| TSH (μIU/mL) | 1.782 (0.924) | 1.727 (0.896) | 1.881 (0.967) | 0.0007 |
| ft4 (ng/dL) | 1.291 (0.161) | 1.314 (0.164) | 1.248 (0.147) | < 0.0001 |
| ft3 (pg/mL) | 3.11 (0.38) | 3.22 (0.35) | 2.91 (0.36) | < 0.0001 |
| Thyroid nodule |  |  |  |  |
| No n (%) | 1226 (66.63%) | 844 (71.10%) | 382 (58.50%) | < 0.0001 |
| Yes n (%) | 614 (33.37%) | 343 (28.90%) | 271 (41.50%) |  |
| < 5.0 mm | 318 (17.28%) | 197 (16.60%) | 121 (18.53%) |  |
| 5.0–9.9 mm | 296 (16.09%) | 146 (12.30%) | 150 (22.97%) |  |
| Isthmus (mm) |  |  |  |  |
| Thickness | 2.3 (0.8) | 2.5 (0.8) | 1.9 (0.7) | < 0.0001 |
| Right lobe (mm) |  |  |  |  |
| Transverse diameter | 15.9 (2.9) | 16.7 (2.8) | 14.4 (2.5) | < 0.0001 |
| Anteroposterior diameter | 13.9 (2.7) | 14.8 (2.5) | 12.2 (2.1) | < 0.0001 |
| Longitudinal diameter | 49.7 (5.5) | 50.2 (5.3) | 48.6 (5.7) | < 0.0001 |
| Left lobe (mm) |  |  |  |  |
| Transverse diameter | 15.5 (2.8) | 16.0 (2.8) | 14.5 (2.3) | < 0.0001 |
| Anteroposterior diameter | 12.1 (2.8) | 13.0 (2.6) | 10.5 (2.4) | < 0.0001 |
| Longitudinal diameter | 47.2 (6.0) | 47.9 (5.7) | 46.0 (6.3) | < 0.0001 |
| Thyroid volume (g) | 10.6 (3.9) | 11.9 (3.8) | 8.3 (2.8) | < 0.0001 |

Continuous variables are expressed as mean (SD). Comparisons between male and female groups were performed using the Student's *t*-test or Fisher's exact test, as appropriate. TSH values were log-transformed for statistical analyses due to their log-normal distribution.

As thyroid nodules measuring 5.0–9.9 mm could potentially affect lobe size in both sexes (Supplementary Table 16), we performed an additional analysis by excluding 296 subjects with nodules in this size range. Multiple regression analysis confirmed that sex remained a significant determinant of thyroid size, independent of height and body weight (Supplementary Table 17). The longitudinal diameter was significantly correlated with age and height. The transverse and anteroposterior diameters were positively correlated with body weight. Thyroid volume was strongly correlated with body weight, but its association with height was no longer significant.

Finally, we calculated reference ranges for thyroid size using data from 1,544 Thyroid-healthy cohort subjects without nodules ≥ 5.0 mm (Table 6). Consistent with the findings in Table 5, the left lobe was smaller than the right lobe in both sexes. For the right lobe, the transverse diameter measured 16.6 ± 2.8 mm in males and 14.1 ± 2.4 mm in females; for the left lobe, it was 15.9 ± 2.8 mm and 14.3 ± 2.2 mm, respectively (p < 0.0001 for males, p = 0.1904 for females). The anteroposterior diameter of the right lobe was 14.7 ± 2.5 mm in males and 12.1 ± 2.1 mm in females, while the left lobe measured 12.9 ± 2.6 mm and 10.4 ± 2.4 mm, respectively (p < 0.0001 for both sexes). The longitudinal diameter of the right lobe was 50.2 ± 5.3 mm in males and 48.5 ± 5.6 mm in females; for the left lobe, 47.9 ± 5.7 mm and 45.7 ± 6.3 mm, respectively (p < 0.0001 for both sexes). Thus, the left lobe was consistently smaller than the right lobe in both sexes, particularly in the anteroposterior and longitudinal diameters.

**Table 6.** Reference ranges for individual thyroid dimensions d by ultrasonography.

|  | Male | Female |
| --- | --- | --- |
| Isthmus (mm) |  |  |
| Thickness | 1.1–4.4 | 0.9–3.8 |
| Right lobe (mm) |  |  |
| Transverse diameter | 11.8–22.8 | 9.7–18.7 |
| Anteroposterior diameter | 10.0–20.5 | 8.1–16.4 |
| Longitudinal diameter | 38.8–59.1 | 37.0–58.1 |
| Left lobe (mm) |  |  |
| Transverse diameter | 10.9–21.7 | 10.3–18.9 |
| Anteroposterior diameter | 7.8–18.2 | 6.0–15.5 |
| Longitudinal diameter | 36.1–58.1 | 33.0–57.5 |
| Thyroid volume (g) | 6.0–20.8 | 4.1–13.7 |

The 2.5th–97.5th percentile was calculated in the Thyroid-healthy cohort, excluding subjects with nodules 5.0–9.9 mm.

## Discussion

In this prospective cohort study conducted in a Japanese medical checkup setting, we comprehensively analyzed data on thyroid examinations, including thyroid function tests, ultrasonography, FDG-PET, and TPOAb measurements. In addition to documenting the prevalence of thyroid dysfunction, we present epidemiological findings. As expected, females were more susceptible to thyroid dysfunction and other abnormalities than males. We also identified factors associated with thyroid function, demonstrating that age, sex, and thyroid volume were dominant determinants. By rigorously excluding subjects with thyroid abnormalities, we defined the Thyroid-healthy cohort and established reference ranges for thyroid size. Furthermore, we identified sex differences and demonstrated that thyroid dimensions correlated with age and body size.

Given that racial differences may affect thyroid function and disease prevalence, our East Asian cohort provides valuable evidence. A meta-analysis of European epidemiological data reported a prevalence of 4.59% for subclinical hypothyroidism, 0.62% for overt hypothyroidism, 0.50% for subclinical hyperthyroidism, and 0.35% for overt hyperthyroidism (11). In the United States, the National Health and Nutrition Examination Survey (NHANES; 2007–2012) found prevalence of 4.3%, 0.33%, 3.20%, and 0.2%, respectively (12). In our cohort, the corresponding values were 2.68%, 0.16%, 2.36% and 0.61%. Another Japanese cohort study of healthy individuals reported an overall prevalence of hypothyroidism of 1.39% (13). These comparisons suggest that hypothyroidism is less common and thyrotoxicosis is more common in Japan than in Europe or the United States. Japan is considered an iodine-sufficient to iodine-excess country (14), and excessive iodine intake is generally associated with an increased risk of developing hypothyroidism (15). In this context, the observed differences are unlikely to be explained by iodine status and may instead reflect other factors, such as racial or genetic differences.

Our cohort also provides epidemiological insights into FDG-PET, ultrasonography, and TPOAb titers. Increased blood flow on ultrasonography was observed in 2.45% of the subjects, with a marked sex difference (1.32% in males vs. 4.14% in females). Schulz *et al.* reported that increased blood flow can occur in hypothyroidism and is strongly associated with elevated serum TSH levels (16). To date, its significance has not been evaluated in population-based cohorts without known thyroid diseases. While ultrasonography is subject to inter-operator variability, the relevance of our findings is supported by their association with thyroid dysfunction, which was more common in subjects with increased blood flow. Furthermore, increased blood flow was associated with TPOAb positivity, which is a well-established risk factor for hypothyroidism. Taken together, in settings where thyroid ultrasonography is readily available, the detection of increased blood flow may facilitate early suspicion of Hashimoto’s disease and hypothyroidism.

While most previous studies have focused on focal thyroid FDG uptake, we specifically examined diffuse FDG uptake to clarify its significance in Hashimoto’s disease and thyroid function. Previous studies have reported diffuse FDG uptake in 3.3% of 1102 cases (5) and 1.8% of 2594 cases (7). In our study, diffuse FDG uptake showed patterns similar to those of increased blood flow, with an overall prevalence of 3.43% (1.48% in males and 6.35% in females), which we report here for the first time. Diffuse FDG uptake was more common in subjects with thyroid dysfunction, particularly overt hypothyroidism, consistent with earlier reports identifying diffuse FDG uptake as a surrogate marker for Hashimoto’s disease (5,6). Moreover, diffuse FDG uptake has been reported as a predictive factor for immune-related adverse events (irAEs) involving the thyroid gland in patients treated with immune checkpoint inhibitors (17,18). Given that female patients are more susceptible to thyroid irAEs (17,19), the high prevalence of diffuse FDG uptake in females may contribute to this increased risk.

A meta-analysis reported a prevalence of thyroid nodules of 24.83% detected on thyroid ultrasonography (20). In our cohort, the prevalence of nodules ≥ 5.0 mm was similar (24.58%). Data on size-specific prevalence are limited; for example, in the Epidemiological Study of Health Effects in Fukushima Emergency Workers, nodules > 5.0 mm were observed in 15.5% of participants (21). The difference from our cohort is likely attributable to sex composition, as the Fukushima study population was predominantly male. In our cohort, the prevalence of nodules ≥ 5.0 mm was lower in males (18.43%) than in females (33.79%). Consistent with previous studies (22–24), we observed a higher prevalence in females and an age-related increase in the frequency of nodules. Moreover, our size-stratified analyses revealed that relatively large nodules were associated with lower TSH levels in females.

TPOAb was detected in 7.19% of the subjects in our cohort, with a higher prevalence in females (10.23%) than in males (5.26%). In NHANES, the prevalence was 11.1% (7.2% in males and 14.8% in females) (12), and in a Japanese health checkup cohort, it was 11.6% (7.2% in males and 15.0% in females) (25). Thus, TPOAb positivity was less frequent in our study, which may reflect differences in assay methodology: NHANES used a different immunoassay and reference range, whereas the Japanese health checkup cohort used a radioimmunoassay kit. Supporting this interpretation, a Chinese study that used the same manufacturer’s assay as ours, with a cut-off of < 34 IU/mL, reported a prevalence of 7.98% (6.12% in males and 9.90% in females) (26), closely matching our results.

Importantly, we identified the clinical relevance of high TPOAb titers. As shown in Table 3, the prevalence of increased blood flow and diffuse FDG uptake elevated in a titer-dependent manner, whereas the prevalence of thyroid dysfunction increased only in subjects with TPOAb titers ≥ 128 IU/mL. Similarly, a previous cohort study found no significant increase in overt hypothyroidism among subjects with TPOAb titers of 35–105 IU/mL, but a marked increase among those with TPOAb titers of 106–350 IU/mL (27). These findings suggest that low-titer TPOAb positivity reflects a relatively mild autoimmune response that may be self-limiting and insufficient to cause thyroid dysfunction. However, given our cross-sectional design, we cannot exclude the possibility that low-titer positivity may predict future thyroid dysfunction. Nonetheless, low-titer TPOAb positivity is unlikely to confer a short-term risk of hypothyroidism.

We further investigated the factors associated with thyroid function, incorporating physiological perspectives. TSH levels were higher in females, whereas fT4 and fT3 levels were higher in males. Although this tendency has been reported in other cohorts (2,28,29), detailed statistical analyses are lacking. To address the potential bias from the higher prevalence of thyroid abnormalities in females, we confirmed sex-related differences in thyroid function using multivariate regression analyses. Age was another major determinant. TSH levels increased with age in both sexes, whereas fT4 and fT3 levels declined with age in males, with minimal associations observed in females. Similar patterns have been reported, with estrogen proposed as a contributing factor (30).

The association between low body weight and low T3 levels is well recognized in non-thyroidal illness, whereas the relationship between obesity and thyroid function remains controversial (31). In our cohort, BMI was positively correlated with fT3 levels in both sexes and TSH levels in females; both associations remained significant in multivariate analyses. As fT3 elevation was not accompanied by changes in fT4 levels, the involvement of iodothyronine deiodinases is likely. Type 2 iodothyronine deiodinase, which exerts a greater influence on circulating fT3 levels than type 1 (32), may mediate this effect, whereas type 3 iodothyronine deiodinase reduces fT3 levels. The clinical importance of type 2 iodothyronine deiodinase is supported by evidence that its pathogenic variant, Thr92Ala, is associated with reduced fT3 levels (33). The observed TSH elevation in females is also noteworthy; one possible mechanism is leptin-mediated stimulation of thyrotropin-releasing hormone secretion, given that females have higher circulating leptin levels (34,35) and obesity increases leptin production (36–38).

Multivariate analyses confirmed that these factors independently determined thyroid function. Abnormal findings on thyroid examinations were also associated with thyroid function, even within the physiological range. However, diffuse FDG uptake and TPOAb titers ≥ 128 IU/mL showed weaker associations than increased blood flow and thyroid volume: diffuse FDG uptake was associated only with fT4, TPOAb titers ≥ 128 IU/mL only with TSH, whereas increased blood flow and thyroid volume were associated with both TSH and fT4 levels. This discrepancy may reflect that ultrasonographic changes indicate structural alterations in the thyroid gland, whereas immune activity detected by FDG-PET or TPOAb positivity alone may be insufficient to cause thyroid dysfunction.

Another strength of the present study is the rigorous assessment of thyroid abnormalities, which enabled us to define the Thyroid-healthy cohort with no abnormal findings on any thyroid examination. Using this cohort, we performed a detailed analysis of thyroid size. First, males in our cohort had larger thyroid glands, independent of age, height, and body weight. Sex differences in thyroid size have been consistently reported (39–41), and our findings reaffirm this observation. Second, we observed a negative correlation between age and the longitudinal diameter. Although age-related changes in thyroid size remain controversial (40–42), this association may explain why we did not detect significant age effects on other parameters (the isthmus thickness, the transverse diameter, and the anteroposterior diameter) and why previous studies focusing on thyroid volume may have failed to detect size changes (43), as thyroid volume is less sensitive to alterations in a single dimension. Third, we identified a positive correlation between height and the longitudinal diameter, whereas body weight was positively correlated with the isthmus thickness and the transverse and anteroposterior diameters. This discrepancy is intriguing and raises the possibility that the thyroid gland may grow predominantly in the longitudinal dimension in proportion to height.

Finally, we established reference ranges for individual thyroid dimensions using ultrasonography. While previous reports have provided regional reference values for thyroid volume (43), to our knowledge, none have reported ranges for each parameter, namely, the isthmus thickness, the transverse diameter, the anteroposterior diameter, and the longitudinal diameter. We determined the reference values using strict criteria, excluding all subjects with any abnormal findings on thyroid examinations or with nodules ≥ 5.0 mm, thereby ensuring a robust reference set. We hope that these sex-specific reference values will serve as a useful tool for evaluating thyroid enlargement in clinical practice.

This study has several limitations. As Japan is an iodine-sufficient country owing to seaweed consumption (44), our results may differ from those in iodine-deficient regions. As our cohort was composed almost entirely of Japanese individuals, racial differences should be considered. Despite the peer review of ultrasonographic parameters, inter-operator variability cannot be completely excluded. Diffuse FDG uptake was assessed qualitatively rather than quantitatively, precluding the establishment of cutoff values. TgAb was not measured, and inclusion of this parameter might have provided additional insights into Hashimoto’s disease, particularly regarding TPOAb/TgAb positivity patterns.

In summary, this prospective cohort study of Japanese adults reported the prevalence of thyroid dysfunction and abnormal findings on thyroid examinations and integrated epidemiological data from ultrasonography, FDG-PET, and TPOAb measurements. We found that TPOAb levels become clinically significant at titers ≥ 128 IU/mL and that age, sex, and thyroid volume are dominant determinants of thyroid function. The Thyroid-healthy cohort, defined through our stringent screening, enabled us to establish sex-specific reference ranges for thyroid dimensions and confirm associations between thyroid size, age, and body size. These findings provide comprehensive epidemiological and physiological evidence in thyroid health and research.

## Data Availability

All data produced in the present work are contained in the manuscript.

## Acknowledgments

We are grateful to the staff of the Preemptive Medicine and Lifestyle Disease Research Center for their assistance in collecting the data analyzed in this study. We also thank the ultrasound technologists for their expertise in performing thyroid ultrasonography and for their careful review of the images.

