## Supplementary Figures 1-4 for "Comprehensive Thyroid Assessment in a Prospective Japanese Cohort: Epidemiology, Determinants, and Ultrasonographic Reference Ranges"

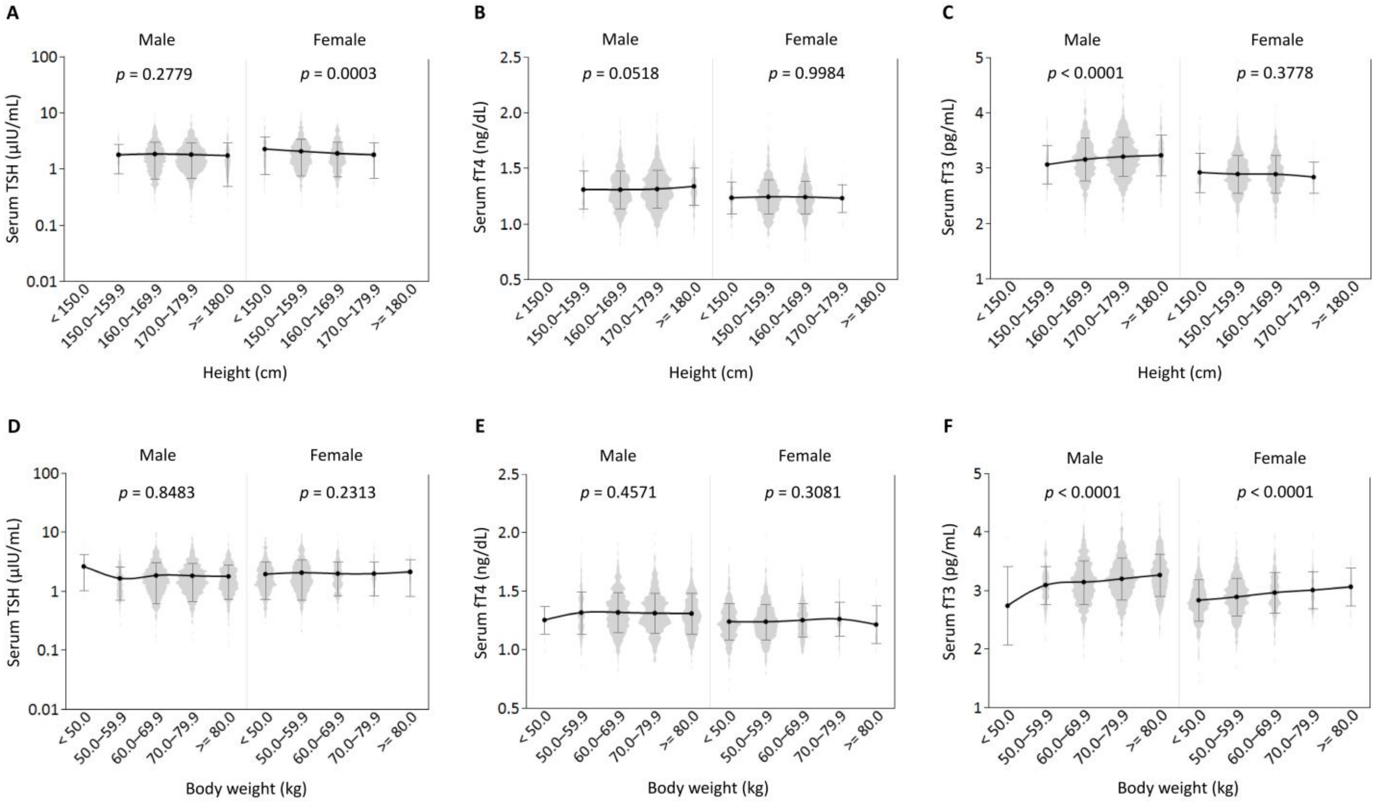


**Supplementary Figure 1.** Association between body size and thyroid function in the Original cohort. (A–C) Trend analyses for height-related changes in thyroid function. Two male subjects shorter than 150.0 cm were excluded due to the small sample size. See Supplementary Table 3 for actual values. (D–F) Trend analyses for body weight-related changes in thyroid function. See Supplementary Table 4 for actual values. Gray violin plots show the distribution of the data, and black circles with connecting lines represent mean ± SD. Log-transformed values were used for statistical analyses of TSH because of its log-normal distribution. Trend analyses were conducted by assigning the median value of each category and modeling it as a continuous variable in regression analyses.

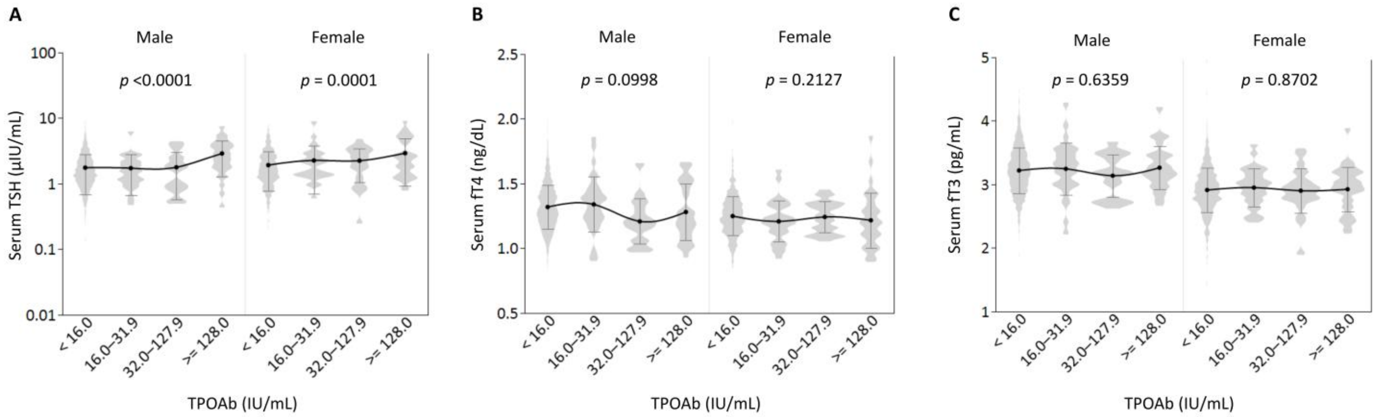


**Supplementary Figure 2.** Trend analyses for TPOAb titer-related changes in thyroid function. See Supplementary Table 11 for actual values. TPOAb, anti-thyroperoxidase antibody. Gray violin plots show the distribution of the data, and black circles with connecting lines represent mean ± SD. Log values were used for statistical analyses of TSH. Trend analyses were performed to determine statistical significance and *p* values.

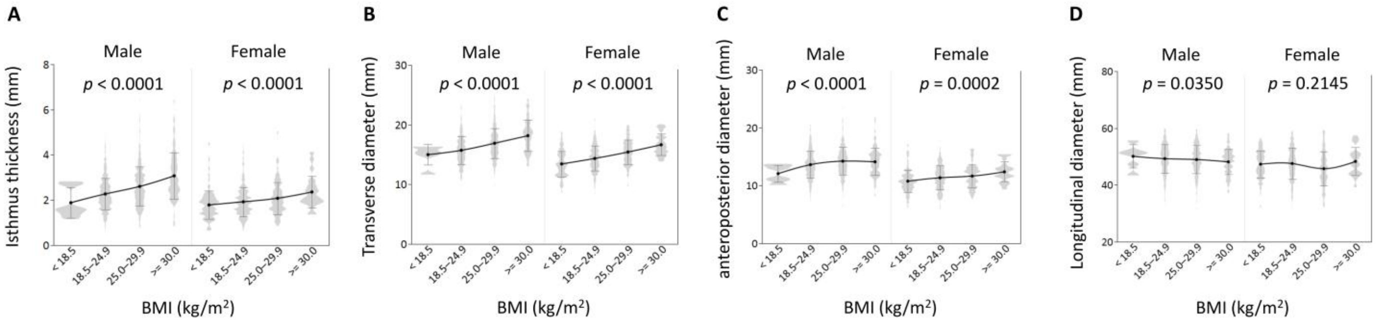


**Supplementary Figure 3.** Trend analyses for BMI-related changes in thyroid size in the Thyroid-healthy cohort. See Supplementary Table 15 for actual values. Gray violin plots show the distribution of the data, and black circles with connecting lines represent mean ± SD. Trend analyses were performed to determine statistical significance and *p* values.

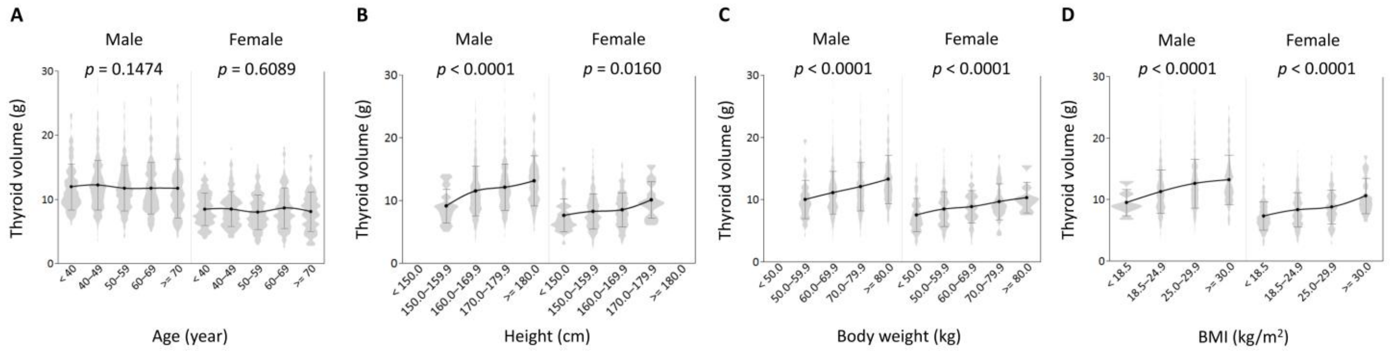


**Supplementary Figure 4.** Association between clinical characteristics and thyroid volume in the Thyroid-healthy cohort. See Supplementary Table 12–15 for actual values. Gray violin plots show the distribution of the data, and black circles with connecting lines represent mean ± SD. Trend analyses were performed to determine statistical significance and *p* values.
